# Seasonal Prediction of Omicron Pandemic

**DOI:** 10.1101/2022.01.13.22269198

**Authors:** Jianping Huang, Yingjie Zhao, Li Zhang, Xu Li, Shuoyuan Gao, Xiaodong Song

## Abstract

The ongoing coronavirus disease 2019 (COVID-19) pandemic has pushed the world in the face of another huge outbreak. In order to have a better understanding on the fast transmission of Omicron variant, we made seasonal predictions on the development of Omicron pandemic globally, as well as 11 key countries. The results demonstrated that the pandemic has an exponential-like growth rate at the initial stage of the outbreak, and will have small resurgences around April and June in north hemisphere countries and south hemisphere countries, respectively.

## Introduction

Different from previous ones, the current outbreak was mainly caused by a newly discovered coronavirus variant which is named as Omicron. There are 6 new coronavirus variants has been categorized as variant of concern by World Health Organization (WHO), namely Alpha, Beta, Gamma, Eta, Delta, and Omicron. Even though all the variants showed the ability of faster spread than the original one, Omicron beat other variants and became the most transmissible variant. Up to now, Omicron variant has spread to more than 120 countries and regions. Some research showed that Omicron variant has the ability of reinfection [1]. The appearance of Omicron variant has subsided the transmission of Delta in South Africa, and it is estimated that more than 75% of the positive tests were infected by Omicron variant [2].

Other than evading from immunity, Omicron has also presented a feature of resistance to antibody [3], and it will likely cause another outbreak worldwide, and become a dominant strain [4]. In this paper, we conducted a seasonal prediction on the spread of Omicron variant by using the Global Prediction System of COVID-19 Pandemic (GPCP) [5,6]. Our prediction results will illustrate the fast spread of Omicron variant, and raise public awareness on it. Moreover, the results can also help governments to make effective measures on containing the pandemic.

## Data and Method

We collected COVID-19 data from the Center for Systems Science and Engineering (CSSE) at Johns Hopkins University [7]. All the data were updated in a daily basis.

The model we used for prediction is a modified SEIR model [8]. Compared with SEIR model, this model added three new coefficients, namely P (protected population), Q (quarantined population), and D (dead population). In addition, we also took temperature and humidity into consideration, and calibrated the transmission rate *β* as *β* = *β*_0_ + *β*_1_*F*(*T*_2*m*_) + *β* _2_*F*(*RH*_2*m*_), where *F*(*T*_2*m*_) and *F*(*RH*_2*m*_) are the probability distribution functions of temperature and humidity obtained by Huang et al. [5], where *T*2*m* represents the temperature at 2m above ground level, and H2*m* is the humidity at 2m above the ground level. *β*_0_ is the original transmission rate of the virus itself, and *β*_1_ and *β*_2_ represent the transmission rates when temperature and humidity are included, respectively. Moreover, the weekly and seasonal cycle of COVID-19, and public behaviors and government policies are parameterized in the GPCP system to make prediction more accurate [9,10]. The model used in the system can be expressed as follow:

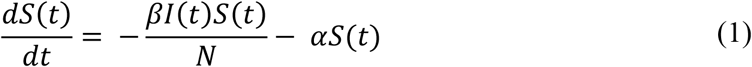

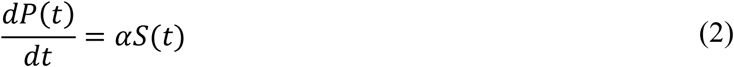

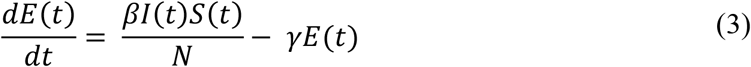

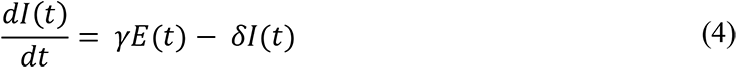

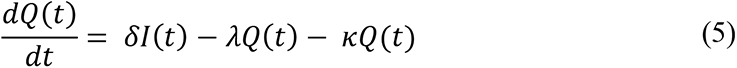

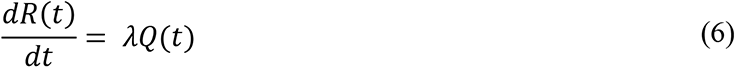

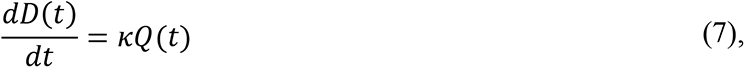

Where S + P + E + I + Q + R + D = N (total population).

## Result

Since Omicron variant was first identified in South Africa on 24th November 2021, it has been spreading to the world at a fast pace. Here, we made prediction on the spread of Omicron variant globally, as well as the spread in some specific countries. The prediction results are shown in Figure 1. From Figure 1, we can see that Omicron exhibits the ability of fast spread, and it is almost spreading at an exponential rate worldwide, and the peak value likely appears around 21st January 2022, with the highest value reach to 308,3988. A small resurgence will appear on 29th April 2022, and the daily confirmed cases will be 183,8134, then the number of daily confirmed cases will decrease, and this trend is consistent with the seasonality of COVID-Moreover, we also selected 11 most affected countries during COVID-19 pandemic, and predict the development trend of the epidemic in each country. The results presented in Figure 1 and Table 1 demonstrate that all countries show exponential-like growth rate at initial stage of the spread of Omicron variant. Even when the south hemisphere countries (Argentina and Australia) are in summer time, they are still suffering from the heavy hit of Omicron variant. Among all the countries, the US is affected by Omicron the most, and the daily confirmed cases will reach to 92,6753 around 18th January 2022. Europe has become the hot-spot for Omicron variant, by the end of June, there will be 39% of the population get infected of COVID-19 in the UK. Other European countries, such as Spain, France, Italy, and Germany will have 77%, 79%, 55%, and 21% of the population get infected with COVID-19, respectively. Due to seasonality of COVID-19, the north hemisphere countries will have small resurgences during spring time, while for south hemisphere countries, the resurgences will occur around July. Apart from the high transmissibility of Omicron variant, the relaxation of control measures, and the large-scale human mobility and gatherings during Christmas holiday are two of the main causes that lead to this huge outbreak of COVID-19 pandemic.

**Table 1:** Total cases from 2022.01 to 2022.06 in each country and the globe

| Country | Total cases (2022.01-06) | Country | Total cases (2022.01-06) |
| --- | --- | --- | --- |
| Global | 366,339,845 | Germany | 10,232,119 |
| US | 99,105,636 | Argentina | 10,934,971 |
| UK | 12,360,502 | Australia | 4,760,583 |
| Spain | 29,079,588 | Turkey | 11,191,154 |
| Italy | 27,798,569 | India | 32,813,076 |
| France | 41,092,489 | Canada | 4,646,398 |

**Figure 1:**
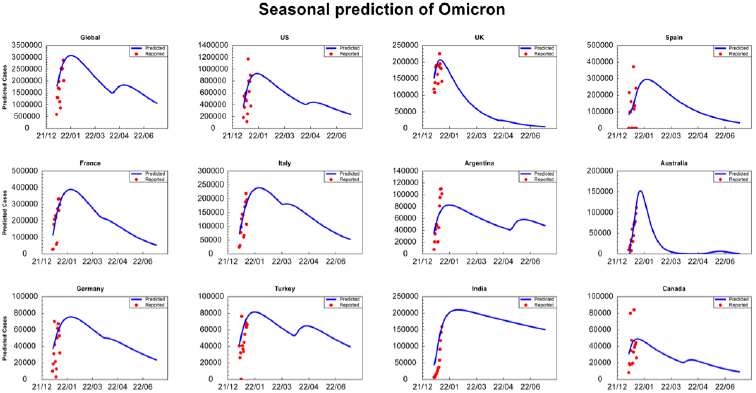
The transmission trend of omicron pandemic in the US, the UK, Spain, France, Italy, Argentina, Australia, Germany, Turkey, India, Canada, and the globe, respectively. The red point represents the real data of daily confirmed cases from 26th December, 2021. The blue curve line shows the predicted daily confirmed cases from 26th December, 2021 to 30th June, 2022.

## Conclusion and Discussion

Omicron variant as a newly discovered coronavirus variant still has many features have not been studied through yet. But the its fast transmission speed is undoubtedly worrying. Unlike previous variants, such as Alpha, Beta, Delta, etc., which can be contained through effective vaccination policy and non-pharmaceutical interventions [11]. Even under current circumstance, that many countries have large population fully vaccinated, Omicron variant still shows an unstoppable spreading trend, and the recent outbreak of COVID-19 is worse than any of that happened before.

Our prediction results showed the destructiveness of Omicron variant and it will continue affecting the whole world for the next 6 months. Before the effectiveness vaccines have been developed, every country should implement strict and effective interventions to slow down the transmission speed of Omicron variant. Especially for countries with undeveloped health systems, if no effective interventions have been employed, the health systems will be overwhelmed, and cause disastrous consequences.

## Data Availability

All data produced in the present work are contained in the manuscript

https://github.com/CSSEGISandData/COVID-19

## Acknowledgments

This work was jointly supported by the National Science Foundation of China (41521004) and the Gansu Provincial Special Fund Project for Guiding Scientific and Technological Innovation and Development (Grant No. 2019ZX-06). The authors acknowledge the Center for Systems Science and Engineering (CSSE) at Johns Hopkins University for providing the COVID-19 data.

